# Quantifying a Minimal Detectable Change within World Rugby’s Head Injury Assessment protocol for elite rugby: a longitudinal observational cohort study

**DOI:** 10.64898/2026.11.12.26367712

**Authors:** James Brown, Ross Tucker, Lindsay Starling, Martin Raftery, Éanna Cian Falvey

## Abstract

**Objective:** Within World Rugby’s Head Injury Assessment (HIA) protocol for elite rugby union, if a player scores one worse than his/her baseline, the sub-test result is deemed abnormal. This analysis aims to quantify year-to-year test-retest reliability of each sub-test in the screens to establish a minimal detectable change (MDC) from the Sports Concussion Assessment Tool (SCAT) baseline testing that is used within the HIA protocol.

**Methods:** SCAT baseline and clinical data are captured by World Rugby’s SCRM App and database for professional men’s and women’s global domestic competitions. Baseline and clinical data were linked on a unique SCRM ID. Test-retest reliability of annual baselines was evaluated using absolute agreement, single-measure intraclass correlation coefficients and standard error of measurement. MDC at a 90% level (MDC_90_) were calculated using a distribution-based approach, comparing players with and without an intervening concussion.

**Results:** Data from 7,686 players (20% women) were analysed, including 2,003 with recorded concussions. Reliability metrics did not differ between players with and without an intervening concussion. Rounded MDC_90_ estimates exceeded one unit for immediate memory, delayed recall and single leg balance, whereas digits backwards, concentration and tandem gait all equalled one unit.

**Conclusion:** These analyses show that annual SCAT baselines in professional rugby union are generally stable across seasons, irrespective of interim concussion. However, the MDC_90_ thresholds of certain SCAT tests suggest that greater Medical Doctor discretion could be allowed when considering if an abnormal test relative to baseline is clinically relevant.

**What is already known:**

- Similar analyses have been conducted within head injury management for ice hockey
- In this previous study, the authors quantified variation from the two most recent Sports Concussion Assessment Tool (SCAT) baselines for all players to calculate a “Reliable Change Index” that could be applied when evaluating if a player had a suspected head injury

**What are the new findings:**

- Using comparable methods to that that were used in professional ice hockey players, this study of elite men and women ice hockey players used all available SCAT baselines to quantify baseline variation, which was termed the “Minimal Detectable Change” (MDC).
- In this, the first study of this nature in professional rugby players, we found that certain SCAT tests, such as the cognitive tests have a high absolute amount of year-year variation, which means that their corresponding MDC would be larger.
- Ignoring this test-specific variation could lead a clinician to believe a player has performed worse than his/her baseline, while this change could actually be within normal variation for that specific test.

## Introduction

World Rugby, the international governing body of rugby union, has a Head Injury Assessment (HIA) protocol in place for professional players [1] . The HIA protocol uses the Sports Concussion Assessment Tool (SCAT), to test players annually at baseline during pre-season [1] . If a player is suspected of having a head injury during the season, the medical team repeat the SCAT on the player at three time points following the head impact, and compare the player to his/her own baseline as part of their clinical assessment.

The SCRM App was developed by World Rugby to assist medical teams in performing the SCAT at baseline and during clinical assessments. This App also records and stores all data captured during this process enabling periodic optimisation of the HIA protocol through research, described previously.[2] .

Professional ice hockey has a comparable process, with annual baseline and clinical screening using the SCAT [3]. A recent analysis assessed test-retest reliability of various SCAT domains within these players [3]. These analyses determined that a 2-point decline in the tandem gait constituted reliable change, or a clinically meaningful decline. By contrast, a one-point decline in this test would indicate change that could be explained by test-retest variation. Similar analyses have been conducted in other fields and this quantification is has been referred to as a minimum detectable change (MDC) [4] .

This study aims to quantify test-retest reliability within the SCAT tests that are used as part of World Rugby’s HIA protocol in professional men and women rugby players.

## Methods

### Study design

This was a longitudinal observational study of routinely collected clinical data.

### Setting

The baseline and HIA data analysed were part of data that are routinely collected as part of World Rugby’s HIA protocol, between 1 January 2023 and 31 December 2025. World Rugby’s HIA assessments are largely based on the latest SCAT tool, SCAT6 [5]. World Rugby’s HIA assessments differ from the SCAT6 in the following ways: there is no double-leg stance or tandem gait balance test; there is no upper limit of errors counted for the balance tests; and for months backwards, there is no time limit. Also, at the first testing time point during the HIA (HIA1), only 11 of the 22 symptoms are asked [1].

Both baseline and HIA data are collected by Medical Doctors who receive specific training in administering the HIA protocol. Baseline data are generally collected during pre-season periods in the medical rooms of the team, but can also be collected before major competitions, resulting in players having more than one baseline per year. Clinical HIA data are captured if the player is suspected of having a concussion and will occur in a dedicated medical room of a stadium.

### Participants

Only baselines that were collected as part of a 15-a-side team were selected by actively excluding all sevens teams. Any player that only had more than one baseline in the period of interest (2023-2025) was included. Finally, if a particular player had more than one complete baseline for the same team/competition within 90 days of another one, then only the most recent baseline was selected for that player in that 90-day period. A complete list of competitions is provided in supplementary table 1.

HIA data were matched with the correct player based on the unique player ID allocated to players by the system. Once baseline and HIA data were matched, a grouping variable indicated if a player had a concussion in the period between baseline collection (January 2023 to December 2025). A concussion diagnosis was made according to World Rugby’s operational definition: the presence of a Criteria 1 sign, or an abnormal HIA2 or HIA3 assessment, or a physician’s diagnosis at any point [1] . There were 21,093 baselines captured from 7,686 unique players. Over the same period, there were 5,120 HIA events that were recorded on 3,674 unique players. All 7,686 players were dichotomised according to whether they had had a concussion in the period of interest or not. A player was in the “no concussion” group if, either (1) the player did not have a corresponding HIA event, or (2) the player had only had HIA events with a final diagnosis of “not concussed”. Players were categorised as “concussed” if they had an HIA event in the period of interest that had a final diagnosis of “concussed”.

### Variables

Concussion in the intervening period was included as an exposure variable.

Six SCAT tests were included as outcome variables for this analysis (Table 1): immediate memory, delayed recall, Digits backwards, Months backwards, single-leg stance balance, tandem stance balance test. For these analyses, Concentration (the sum of Digits backwards and Months backwards) was analysed – five outcome variables in total.

**Table 1.**
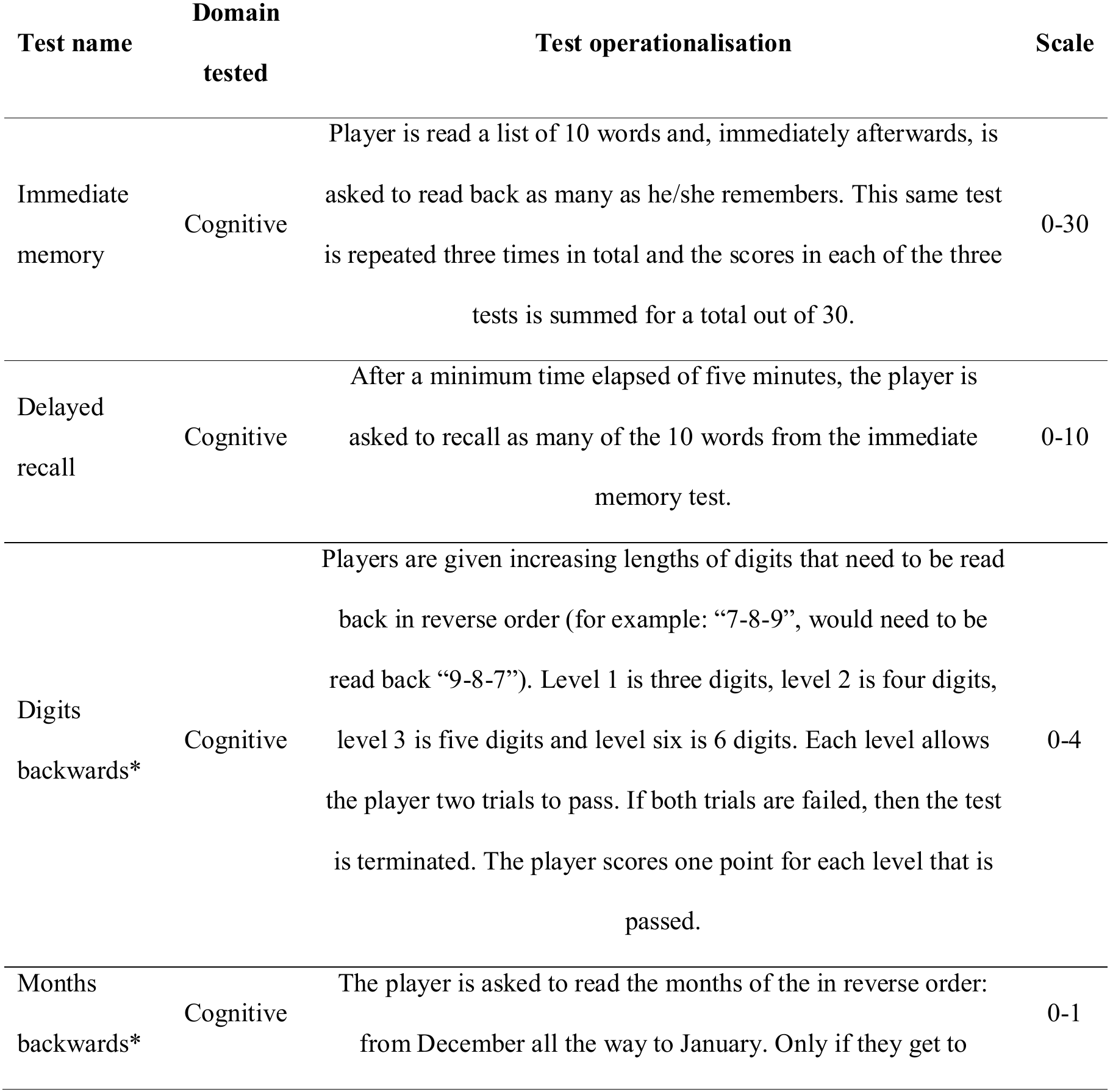
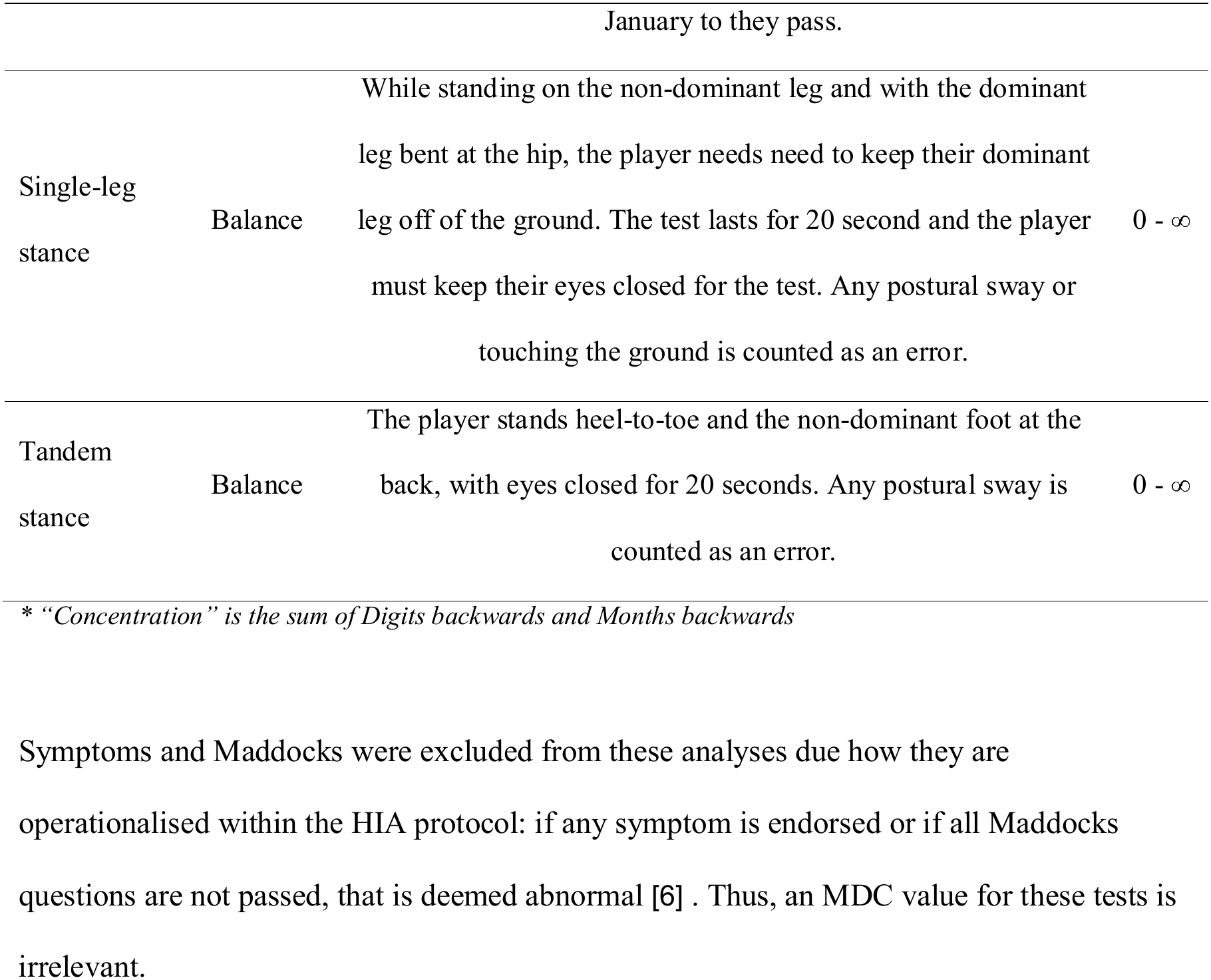
The Sports Concussion Assessment Tool (SCAT) variables that were included as outcomes. All of these variables were captured as part of World Rugby’s baseline tests, within the HIA protocol.

| Test name | Domain tested | Test operationalisation | Scale |
|---|---|---|---|
| Immediate memory | Cognitive | Player is read a list of 10 words and, immediately afterwards, is asked to read back as many as he/she remembers. This same test is repeated three times in total and the scores in each of the three tests is summed for a total out of 30. | 0-30 |
| Delayed recall | Cognitive | After a minimum time elapsed of five minutes, the player is asked to recall as many of the 10 words from the immediate memory test. | 0-10 |
| Digits backwards* | Cognitive | Players are given increasing lengths of digits that need to be read back in reverse order (for example: “7-8-9”, would need to be read back “9-8-7”). Level 1 is three digits, level 2 is four digits, level 3 is five digits and level six is 6 digits. Each level allows the player two trials to pass. If both trials are failed, then the test is terminated. The player scores one point for each level that is passed. | 0-4 |
| Months backwards* | Cognitive | The player is asked to read the months of the in reverse order: from December all the way to January. Only if they get to January to they pass. | 0-1 |
| Single-leg stance | Balance | While standing on the non-dominant leg and with the dominant leg bent at the hip, the player needs need to keep their dominant leg off of the ground. The test lasts for 20 second and the player must keep their eyes closed for the test. Any postural sway or touching the ground is counted as an error. | 0 - ∞ |
| Tandem stance | Balance | The player stands heel-to-toe and the non-dominant foot at the back, with eyes closed for 20 seconds. Any postural sway is counted as an error. | 0 - ∞ |

\* *“Concentration” is the sum of Digits backwards and Months backwards*

Symptoms and Maddocks were excluded from these analyses due how they are operationalised within the HIA protocol: if any symptom is endorsed or if all Maddocks questions are not passed, that is deemed abnormal [6] . Thus, an MDC value for these tests is irrelevant.

### Data sources/measurement

All baseline and clinical data are captured on the SCRM App by Medical Doctors. When performing an HIA assessment, the SCRM App automatically compares it to the player’s own most recent baseline. If the player has performed any worse during the clinical assessments following head impacts than during the relevant prior baseline, the sub-test result is flagged to the Medical Doctor as abnormal. For example, if a player scored 23 out of 30 for their Immediate Memory test during their baseline, a score of 22 or fewer for the Immediate Memory test in their HIA is currently deemed abnormal. Within certain guidelines that were developed empirically by World Rugby [6] , the Medical Doctor administering the test is permitted to overrule test abnormalities [6].

### Bias

We assumed that each baseline score represents a player’s best performance. However, there is no way to control for the baseline or HIA testing conditions. Within the clinical data, clinical judgement determines the diagnosis of concussion, as is the case for all sports-related concussions [5] .

### Study size

No *a priori* sample size calculation was performed as all eligible baselines and HIA events that were recorded during the study period were included.

### Statistical methods

Reliability across repeated annual baselines was quantified with a two-way, mixed-effects, absolute-agreement, single-measure intraclass correlation coefficient (ICC), with 95% confidence intervals (CI) [7]. This form of ICC considers practice effects as part of the error term that is calculated [7]. Variance components were estimated from mixed-effects models with a random intercept for player (player unique ID). For the four cognitive outcomes (Table 1) with more continuous outcomes, a linear mixed-effects model was used. For the two balance tests, that have number of errors as the outcome (Table 1), a Poisson distribution was used [8].

To calculate the Minimal Detectable Change (MDC), we first calculated the standard error of measurement (SEM) from the ICC and the between-assessment standard deviation (SD) [4] .

Finally, the MDC was calculated at the arbitrarily chosen 90% level with a distribution-based approach [4,9]:

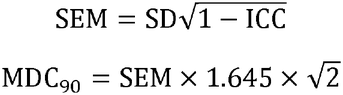

The value of 1.645 bounds 90% of the distribution of change scores expected from measurement error alone, with 5% in each distribution tail. MDCs were rounded to the nearest whole number, as was done in ice hockey [3], because all sub-tests are evaluated in whole units within the HIA protocol. Under the current (‘status quo’) HIA criterion, any decline (change <0) is classified as abnormal [1] . Once this MDC is calculated, a test would only be considered abnormal if the decline is equal to or greater than the MDC. If a test has a rounded MDC score of one unit, then there would be no difference between current and MDC criteria. The implications of these MDCs were assessed by examining the HIA tests that were administered clinically during rugby matches (known as HIA1 assessments) during the period of interest. The proportion of abnormalities that was calculated and compared the proportion of abnormalities that would be present once the MDC was considered.

As a form of “sensitivity analysis”, a MDC table was also calculated at the 80% level (supplementary Table 2) to assess the implications of a different level on the rounded MDC values.

ICC, SEM and MDC were calculated for the entire cohort and separately for the concussion and no-concussion groups. Differences between concussed and not-concussed groups were not formally tested statistically as the most important outcome was the MDC rounded value for each group, not whether the groups were statistically different.

Some specific tests had missing data that, after investigation, was decided to be missing at random. For example, if a player already has a valid and complete baseline in the system, the Medical Doctor conducting a new baseline can choose to skip optional test, such as immediate memory or the balance tests. Thus, the number of valid tests that were used to calculate each value are described in the Results section.

All analyses were performed in R (version 2026.10, Posit Software). Intraclass correlation coefficients (ICCs) and standard errors of measurement (SEMs) were calculated using the **psych** package [10] .

### Data access and cleaning

JB performed spot-checks of the original (raw) and de-identified data, as well as targeted spot-checks on any potentially implausible data that were identified through initial data cleaning and checking.

## Results

In total, there were 7,686 unique players that had two or more baselines recorded between 2023 and 2025 (Table 2) and were thus considered for these analyses. These 7,686 players had a total of 21,093 baseline assessments between them, an average of nearly 3 baselines, per player. The majority of the 7,686 players were men (6,164, 80%) and the majority of players (n = 4,477, 74%) in the analysis had not had a concussion in the period (“no intervening concussion”) (Table 2).

**Table 2.** Cohort description.

|  | **Unique players** |  |  | **Total baseline assessments** |  |  |
|---|---|---|---|---|---|---|
|  | **Men** | **Women** | **Total** | **Men** | **Women** | **Total** |
| Entire cohort | 6,164 | 1,522 | 7,686 | 16,712 | 4,381 | 21,093 |
| Intervening concussion | 1,687 | 316 | 2,003 | 5,006 | 1,074 | 6,080 |
| No intervening concussion | 4,477 | 1,206 | 5,683 | 11,706 | 3,307 | 15,013 |

The rounded MDC_90_ value for Digits backwards, Concentration and Tandem stance were all one unit (Table 3). The rounded MDC_90_ value for immediate memory was four units, and it was two units for delayed recall and single-leg stance. For all tests, the rounded MDC_90_ values were the same for the group of players that had had a concussion in the intervening period (“Intervening concussion”) as those who had not had a concussion (“No intervening concussion”) during the same period. The rounded MDC_90_ values were also not different between men and women (Supplementary Table 3).

**Table 3.** Intraclass correlation coefficient (ICC), standard error of measurement (SEM) and Minimal Detectable Change (MDC) at the 90% level (MDC_90_). Values are shown separately for those who did and didn’t have a concussion in between baselines.

| Test | Cohort | Valid tests | ICC | ICC 95% CI | SEM | MDC<sub>90</sub> | Rounded MDC<sub>90</sub> |
|---|---|---|---|---|---|---|---|
| Immediate Memory | Entire Cohort | 16,612 | 0.743 | [0.734, 0.753] | 1.668 | 3.881 | 4 |
|  | Intervening Concussion | 4,669 | 0.731 | [0.712, 0.749] | 1.685 | 3.919 | 4 |
|  | No intervening Concussion | 11,943 | 0.747 | [0.736, 0.758] | 1.661 | 3.863 | 4 |
| Delayed Recall | Entire Cohort | 16,612 | 0.766 | [0.757, 0.775] | 0.813 | 1.891 | 2 |
|  | Intervening Concussion | 4,669 | 0.754 | [0.737, 0.771] | 0.823 | 1.915 | 2 |
|  | No intervening Concussion | 11,943 | 0.771 | [0.761, 0.781] | 0.808 | 1.880 | 2 |
| Digits Backwards | Entire Cohort | 21,090 | 0.694 | [0.684, 0.704] | 0.446 | 1.037 | 1 |
|  | Intervening Concussion | 6,080 | 0.678 | [0.659, 0.697] | 0.442 | 1.027 | 1 |
|  | No intervening Concussion | 15,010 | 0.698 | [0.686, 0.709] | 0.448 | 1.042 | 1 |
| Concentration | Entire Cohort | 21,087 | 0.688 | [0.678, 0.698] | 0.465 | 1.082 | 1 |
|  | Intervening Concussion | 6,080 | 0.674 | [0.655, 0.693] | 0.461 | 1.072 | 1 |
|  | No intervening Concussion | 15,007 | 0.691 | [0.680, 0.703] | 0.467 | 1.086 | 1 |
| Single-leg stance | Entire Cohort | 21,091 | 0.652 | [0.641, 0.663] | 0.935 | 2.174 | 2 |
|  | Intervening Concussion | 6,080 | 0.666 | [0.645, 0.687] | 0.913 | 2.125 | 2 |
|  | No intervening Concussion | 15,011 | 0.646 | [0.633, 0.659] | 0.943 | 2.193 | 2 |
| Tandem stance | Entire Cohort | 21,091 | 0.806 | [0.800, 0.812] | 0.461 | 1.073 | 1 |
|  | Intervening Concussion | 6,080 | 0.815 | [0.804, 0.827] | 0.428 | 0.996 | 1 |
|  | No intervening Concussion | 15011 | 0.802 | [0.795, 0.810] | 0.474 | 1.104 | 1 |

*95% CI: 95% confidence interval*

The sensitivity analysis that calculated MDC at the 80% level is shown in supplementary table 2. In summary, MDC_80_ is different to the MDC_90_ rounded value for the following tests: immediate memory (3 units for MDC80 instead of 4 for MDC90) and delayed recall (1 unit for MDC80 instead of 2 for MDC90). The remaining tests (digits backwards, concentration, single-leg and tandem stance balance tests) produced the same MDC at 80% and 90%.

In an attempt to quantity the impact that the application of these MDC_90_ would have on the clinical testing of players, we examined 4,264 HIA assessments (“HIA1” assessments) in the period of interest (figures 1-3). This was only performed for the three tests that had an MDC_90_ greater than one unit, as sub-tests with an MDC_90_ of one unit are unaffected by any change in application.

**Figure 1.**
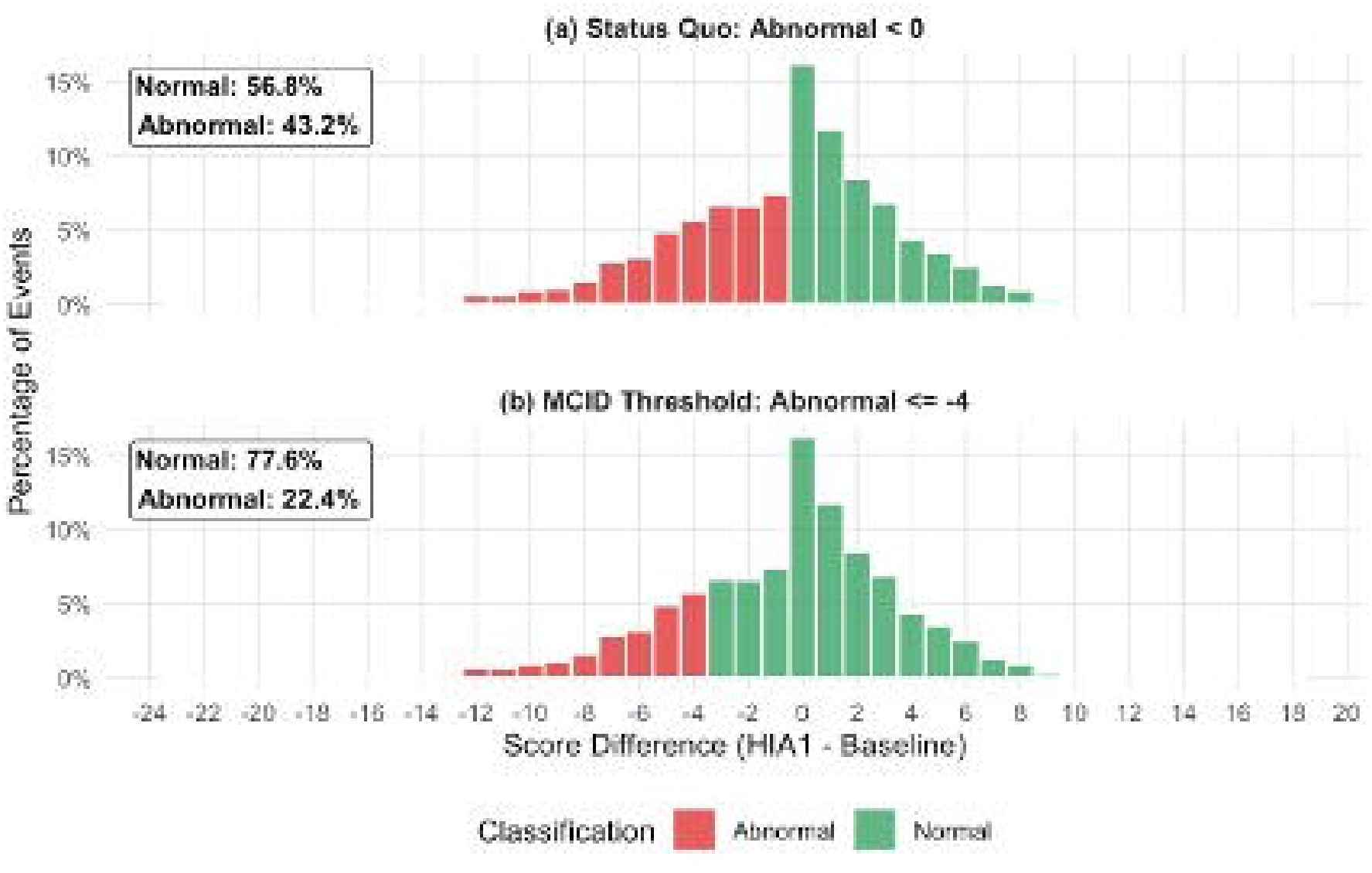
Immediate memory test, scored out of 30. (a) HIA1 assessments currently classified as abnormal by status quo, and (b) if an MDC_90_ (4 units) had been considered.

For immediate memory, 43% of HIA1 assessments are currently flagged as abnormal (worse by one more relative to baseline (Figure 1a)). The application of the MDC_90_ of four units means that 22% of assessments would be abnormal for immediately memory (figure 1b).

For delayed recall, 41% of HIA1 assessments are flagged as abnormal under the status quo for being one score or more below baseline (Figure 2a). Had an MDC_90_ (2 units) been applied to this test, the proportion of abnormal delayed recall scores would drop by 13%, to 28% of assessments (figure 2b).

**Figure 2.**
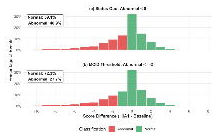
Delayed recall test, scored out of 10. (a) HIA1 assessments currently classified as abnormal by status quo, and (b) abnormal assessments if an MDC_90_ (2 units) were applied.

**Figure 3.**
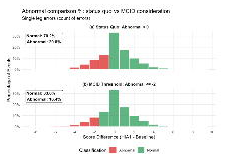
Single-leg balance test, count of errors (a) HIA1 assessments currently classified as abnormal by status quo, and (b) if an MDC_90_ had been considered.

For single-leg errors, 30% of HIA1 assessments are currently flagged as abnormal for producing more errors than in baseline (Figure 3a). The application of an MDC_90_ (2 units) reduces this to 16%, a reduction of 14% points (figure 3b).

## Discussion

This study aimed to quantify baseline test reliability through the minimum detectable change (MDC) of the SCAT tests that are used within World Rugby’s HIA protocol in professional men and women rugby players. We found that the rounded MDC_90_ value for Digits backwards, Concentration and Tandem stance were all one unit (Table 3). This means that any score worse than baseline in these sub-tests is considered abnormal, and is consistent with current use. However, the MDC_90_ value for immediate memory, delayed recall and single-leg stance were greater than one unit, which differs from existing use. The rounded MDC_90_ value for immediate memory was four units, and for both delayed recall and single-leg stance it was two units. The main implication of this finding is that the proportion and number of abnormal sub-tests would decrease with the application of MDC. While four units for immediate memory might initially appear to be a large MDC, it is actually only 13% of the maximum score (30). Relatively, the MDC of 2 units for delayed recall represents a larger percentage (20%) of the test maximum score (10).

The rounded reliable change index (RCI) score for concentration and digits backwards was two units at both the 80% and 90% level in professional ice hockey players study [3]. This is one unit more than in the present study in rugby players. These differences in MDC or RCI are due to the ICC of the ice hockey study being lower (less reliable) than in this rugby cohort. Specifically, the ICC of concentration score in English-speaking ice hockey players was 0.56 (no confidence intervals provided) [3] , while our rugby playing cohort had an ICC of 0.69 (95% CIs: 0.68 to 0.70). The ICC for digits backwards in the ice hockey players was 0.54 (no confidence intervals provided), while our rugby cohort had an ICC of 0.69 (0.68 to 0.70) for the same test. However, there are notable differences between the ice hockey study’s and the present study’s methods. Firstly, the ice-hockey study strictly adheres to the SCAT6 version, as mentioned in the Methods section. There are also differences in how test re-test reliability were quantified between the two tests. Specifically, the ice hockey study only used the last two baselines for a player. It also included a Pearson’s R correlation to determine practice effects, an ICC to quantify reliability in these two baselines and then a formula to calculate RCI. While the formula to calculate the RCI is mathematically very similar to the present study’s quantification of MDC, we used a mixed-effects model, which is able to include multiple baselines in the quantification of baseline-to-baseline variability. This approach has the advantage of including practice/learning effects within the error term of the ICC calculation, and is also able to handle more than two baselines per player. We prioritised inclusion of more than two baselines per player as it would more accurately reflect a player’s longitudinal baseline performance, and reflect the reality of the professional rugby cohort performing multiple baselines in a calendar year. There were no other comparable SCAT tests between the ice hockey and present study.

Various composite components of the SCAT have been assessed for test re-test reliability in college athletes [11]. An ICC of 0.33 (95% CI: 0 – 0.61) was calculated for the Standardised Assessment of Concussion (SAC), including Immediate memory, delayed recall, concentration and orientation. In the present study, the ICCs of the SAC components that were measured (orientation was not included for aforementioned reasons) were far higher and ranged from 0.69 to 0.77 (range of 95% CIs across all: 0.68 – 0.78). This suggests the professional rugby cohort has more stable baselines than in the college students [11], perhaps expected in professional athletes.

It is important to emphasise that any measurement of MDC or RCI is simply a quantification of baseline-to-baseline variability within that cohort [4] . The application of MDCs is not intended to override clinical judgement or opinion, especially for a diagnosis of concussion that requires clinical decision-making [5] . There are also important limitations of this analysis that must be acknowledged. Firstly, as the data were analysed over a relatively short period of time 2023 to 2025, these analyses would need to be repeated periodically, prospectively ensuring validity. The volume of baseline data outlined here assures us that these findings are applicable to the current elite rugby players. We also employed an *a priori* selection criteria for the baselines that were to be included. These decisions were made to reflect clinical reality of professional rugby and the HIA environment as closely as possible. While every scenario cannot be accounted for, we did include a sensitivity analysis for the chosen MDC level, as this was selected arbitrarily (supplementary table 2). The implications, if a 80% level were chosen instead of 90%, are small (3 units instead of 4 for immediate and 1 unit instead of 2 for delayed recall).

If these MDCs were implemented in the HIA protocol, it could potentially reduce the number of abnormal screen outcomes where the calculated MDC_90_ is greater than one unit. A test that would previously be identified as abnormal, and require clinical judgment from the physician to overrule would now be identified as within MDC limits, and would be less likely to be treated as an abnormal result. The impact of this is to hypothetically reduce abnormal sub-test results by between 13% and 21% points for immediate memory, delayed recall and single-leg balance (figures 1-3). However, this would not negate the need for the practitioner administering the test to apply judgment to decide if the test result is clinically relevant or not, within the existing guidelines [1,6]. This means that it is difficult to accurately forecast the more important outcome of introducing the MDC, which is the proportion of abnormal tests where the clinical result is worse than baseline but not by more than MDC, and which practitioners consider to be clinically relevant. For example, if the MDC is integrated into the

HIA, practitioners may find it more difficult to exercise clinical judgement when an immediate memory score drops by three units rather than just one. This highlights the importance of continuous monitoring of clinical decisions in the HIA, should any change such as the MDC be implemented.

In conclusion, this study quantified test re-test reliability of serial baselines in elite men and women rugby players. This reliability and variability was packaged as an MDC so that it could be implemented within the HIA protocol. The rounded MDC_90_ for immediate memory was four units, and it was two units for both delayed recall and single-leg stance. If this MDC was operationalised within the HIA, it means that practitioners would be better guided to understand if a change in a sub-test score is clinically relevant or the result of random variation. Currently, any sub-test outcome worse than baseline is highlighted as abnormal to the practitioners. The other tests that were quantified (concentration and tandem stance) would be unchanged in how they are operationalised as their calculated MDC was one unit.

## Data Availability

The sharing of these data was not covered by aforementioned consent process, but the corresponding author can be contacted to request a copy of the de-identified data and each request will be considered. The analysis code can be shared upon request.

## Ethics approval statement and patient consent

Ethics approval was received from World Rugby’s Internal Ethics Committee. All players provided written informed consent for data gathered as part of the World Rugby Concussion management programme to be used for research in a de-identified manner. This informed consent is captured at the start of each season.

