## Supplementary Tables (1-3) for "Quantifying a Minimal Detectable Change within World Rugby’s Head Injury Assessment protocol for elite rugby: a longitudinal observational cohort study"

### Supplementary files

**Supplementary table 1. Assessments counted by competitions that the team was included in at the time of collection.**

| **Tournament name** | **Men** | **Women** | **Total** |
| --- | --- | --- | --- |
| 2025 British & Irish Lions Tour (AUS) | 124 | 0 | 124 |
| 2025 Men's International | 350 | 0 | 350 |
| 2025 PWR Cup | 0 | 791 | 791 |
| 2025 Women's International | 0 | 7 | 7 |
| 2026 Men's HIA Games | 91 | 0 | 91 |
| 2026 U20 Six Nations | 17 | 0 | 17 |
| 2026 Womens Six Nations | 0 | 11 | 11 |
| 4N SAR 2023 | 1 | 0 | 1 |
| 6 Nations Women's Summer Series 2025 | 0 | 106 | 106 |
| Africa Women's XV Cup 2024 | 0 | 24 | 24 |
| Allianz Premier 15s 22/23 | 0 | 4 | 4 |
| Asia Rugby Emirates Men's Championship 2025 | 4 | 0 | 4 |
| Autumn Nations Series 2025 | 21 | 0 | 21 |
| Celtic Challenge 2026 | 0 | 74 | 74 |
| Celtic Challenge 24/25 | 0 | 26 | 26 |
| Challenger Series Men's 2023 | 4 | 0 | 4 |
| Challenger Series Women's 2023 | 0 | 2 | 2 |
| Currie Cup 2023 | 12 | 0 | 12 |
| Currie Cup 2024 | 86 | 0 | 86 |
| Currie Cup 2025 | 235 | 0 | 235 |
| EPCR Challenge Cup 2026 | 7 | 0 | 7 |
| EPCR Challenge Cup 23/24 | 506 | 0 | 506 |
| England Championship League 24/25 | 63 | 0 | 63 |
| England PWR League 2024-25 | 0 | 68 | 68 |
| England Premiership Championship Cup 23/24 | 281 | 0 | 281 |
| England Premiership Cup 2024-25 | 157 | 0 | 157 |
| England Premiership League 23/24 | 541 | 0 | 541 |
| England Premiership League 24/25 | 71 | 0 | 71 |
| England Women's Premiership Cup 23/24 | 0 | 453 | 453 |
| English Championship 2026 | 400 | 0 | 400 |
| FFR Access Match 2025 | 75 | 0 | 75 |
| FFR/LNR HIA Matches 2024 | 25 | 0 | 25 |
| Farah Palmer Cup 2025 | 0 | 230 | 230 |
| Heineken Champions Cup 22/23 | 4 | 0 | 4 |
| Investec Champions Cup 23/24 | 14 | 0 | 14 |
| Investec Champions Cup 24/25 | 1278 | 0 | 1278 |
| Japan League 2 2026 | 409 | 0 | 409 |
| Japan League One 2026 | 536 | 0 | 536 |
| Japan League One Div 1 23/24 | 337 | 0 | 337 |
| Japan League One Div 1 24/25 | 454 | 0 | 454 |
| Japan League One Div 2 23/24 | 142 | 0 | 142 |
| Japan League One Div 2 24/25 | 412 | 0 | 412 |
| Japan League One Div1 vs Div2 Play-off 2 23/24 | 233 | 0 | 233 |
| Junior Men’s Internationals 23 | 48 | 0 | 48 |
| League One 22/23 | 12 | 0 | 12 |
| MLR 2023 | 230 | 0 | 230 |
| MLR 2024 | 543 | 0 | 543 |
| MLR 2025 | 377 | 0 | 377 |
| Melrose Cup Invitational 2026 - Womens | 0 | 5 | 5 |
| Men's Friendly HIA Games 2024 | 612 | 0 | 612 |
| Men's HIA Games 2025 | 597 | 0 | 597 |
| Men's International 2024 | 864 | 0 | 864 |
| Mens Internationals 22/23 | 46 | 0 | 46 |
| Mens Rugby World Cup 2027 Qualifying | 23 | 0 | 23 |
| New Zealand FPC 2023 | 0 | 240 | 240 |
| New Zealand FPC 2024 | 0 | 297 | 297 |
| New Zealand NPC 2023 | 156 | 0 | 156 |
| New Zealand NPC 2024 | 155 | 0 | 155 |
| New Zealand NPC 2025 | 123 | 0 | 123 |
| OCEANIA WOMEN’S CHAMPIONSHIP 2024 | 0 | 6 | 6 |
| Oceania U20s Challenge 2025 | 8 | 0 | 8 |
| Oceania Women's XV Championships 2025 | 0 | 17 | 17 |
| PACIFIC FOUR SERIES 2024 | 0 | 48 | 48 |
| PWR 2026 | 0 | 103 | 103 |
| PWR Cup 2025 | 0 | 166 | 166 |
| Pacific Nations Cup 2025 | 67 | 0 | 67 |
| Premiership Rugby Cup 2026 | 764 | 0 | 764 |
| Pro D2 2023-24 | 333 | 0 | 333 |
| Pro D2 2024-25 | 327 | 0 | 327 |
| Pro D2 2026 | 482 | 0 | 482 |
| RAN Mens Olympic qualifiers 2023 | 2 | 0 | 2 |
| RAN Womens Olympic qualifiers 2023 | 0 | 4 | 4 |
| RUGBY EUROPE INTERNATIONAL CHAMPIONSHIP 2025 | 83 | 0 | 83 |
| Rugby Africa Cup 2025 | 107 | 0 | 107 |
| Rugby Championship U20 2025 | 53 | 0 | 53 |
| Rugby Europe Championship 2023 | 10 | 0 | 10 |
| Rugby Europe Championship 2024 | 63 | 0 | 63 |
| Rugby Europe Men's Championship 2026 | 5 | 0 | 5 |
| Rugby Europe Women 2025 | 0 | 28 | 28 |
| Rugby Europe Women's Championship 2024 | 0 | 17 | 17 |
| Rugby World Cup 2023 | 85 | 0 | 85 |
| Rugby World Cup 2027 Final Qualification Tournament | 57 | 0 | 57 |
| SANZAAR Under 20s | 99 | 0 | 99 |
| Six Nations 2025 | 3 | 0 | 3 |
| Six Nations U20 2024 | 161 | 0 | 161 |
| Six Nations U20 2025 | 73 | 0 | 73 |
| Super Rugby Americas 2024 | 158 | 0 | 158 |
| Super Rugby Americas 2025 | 175 | 0 | 175 |
| Super Rugby Americas 2026 | 1 | 0 | 1 |
| Super Rugby Aupiki 2023 | 0 | 35 | 35 |
| Super Rugby Aupiki 2024 | 0 | 109 | 109 |
| Super Rugby Aupiki 2025 | 0 | 42 | 42 |
| Super Rugby Pacific 2023 | 42 | 0 | 42 |
| Super Rugby Pacific 2024 | 135 | 0 | 135 |
| Super Rugby Pacific 2025 | 135 | 0 | 135 |
| Super Rugby Pacific 2026 | 51 | 0 | 51 |
| Super W 2023 | 0 | 34 | 34 |
| Super W 2024 | 0 | 158 | 158 |
| Super W 2025 | 0 | 93 | 93 |
| Top 14 2023-24 | 5 | 0 | 5 |
| Top 14 2024-25 | 346 | 0 | 346 |
| Top 14 2026 | 454 | 0 | 454 |
| U20 Championship 2023 | 46 | 0 | 46 |
| U20 Championship 2024 | 138 | 0 | 138 |
| U20 Championship 2025 | 94 | 0 | 94 |
| U20 Trophy 2023 | 16 | 0 | 16 |
| U20 Trophy 2024 | 63 | 0 | 63 |
| URC 2023-24 | 30 | 0 | 30 |
| URC 2024-25 | 774 | 0 | 774 |
| URC 2026 | 551 | 0 | 551 |
| WORLD RUGBY PACIFIC CHALLENGE 2024 | 39 | 0 | 39 |
| WXV 2 2023 | 0 | 141 | 141 |
| WXV 3 2023 | 0 | 53 | 53 |
| WXV 3 2024 | 0 | 196 | 196 |
| Women's International 2024 | 0 | 49 | 49 |
| Women's Internationals 23 | 0 | 133 | 133 |
| Women's Rugby World Cup 2025 | 0 | 491 | 491 |
| Women's U20 Six Nations 2024 | 0 | 63 | 63 |
| Womens Six Nations 2024 | 0 | 56 | 56 |
| World Rugby Junior World Championship 2026 | 1 | 0 | 1 |
| World Rugby Pacific Four Series 2026 | 0 | 1 | 1 |
| **Total** | **16,712** | **4,381** | **21,093** |

**Supplementary Table 2. Intraclass correlation coefficient (ICC), standard error of measurement (SEM) and Minimal Detectable Change (MDC) at the 90% level (MDC_90_). Values are shown separately for those who did and didn’t have a concussion in between baselines.**

| **Test** | **Cohort** | **Valid tests** | **ICC** | **ICC 95% CI** | **SEM** | **MDC_80_** | **Rounded MDC_80_** |
| --- | --- | --- | --- | --- | --- | --- | --- |
| Immediate Memory | Entire Cohort | 16,612 | 0.743 | [0.734, 0.753] | 1.668 | 3.024 | 3 |
|  | Intervening Concussion | 4,669 | 0.731 | [0.712, 0.749] | 1.685 | 3.054 | 3 |
|  | No intervening Concussion | 11,943 | 0.747 | [0.736, 0.758] | 1.661 | 3.011 | 3 |
| Delayed Recall | Entire Cohort | 16,612 | 0.766 | [0.757, 0.775] | 0.813 | 1.474 | 1 |
|  | Intervening Concussion | 4,669 | 0.754 | [0.737, 0.771] | 0.823 | 1.492 | 1 |
|  | No intervening Concussion | 11,943 | 0.771 | [0.761, 0.781] | 0.808 | 1.465 | 1 |
| Digits Backwards | Entire Cohort | 21,090 | 0.694 | [0.684, 0.704] | 0.446 | 0.809 | 1 |
|  | Intervening Concussion | 6,080 | 0.678 | [0.659, 0.697] | 0.442 | 0.801 | 1 |
|  | No intervening Concussion | 15,010 | 0.698 | [0.686, 0.709] | 0.448 | 0.812 | 1 |
| Concentration | Entire Cohort | 21,087 | 0.688 | [0.678, 0.698] | 0.465 | 0.843 | 1 |
|  | Intervening Concussion | 6,080 | 0.674 | [0.655, 0.693] | 0.461 | 0.836 | 1 |
|  | No intervening Concussion | 15,007 | 0.691 | [0.68, 0.703] | 0.467 | 0.846 | 1 |
| Single-leg stance | Entire Cohort | 21,091 | 0.652 | [0.641, 0.663] | 0.935 | 1.694 | 2 |
|  | Intervening Concussion | 6,080 | 0.666 | [0.645, 0.687] | 0.913 | 1.656 | 2 |
|  | No intervening Concussion | 15,011 | 0.646 | [0.633, 0.659] | 0.943 | 1.709 | 2 |
| Tandem stance | Entire Cohort | 21,091 | 0.806 | [0.8, 0.812] | 0.461 | 0.836 | 1 |
|  | Intervening Concussion | 6,080 | 0.815 | [0.804, 0.827] | 0.428 | 0.776 | 1 |
|  | No intervening Concussion | 15,011 | 0.802 | [0.795, 0.81] | 0.474 | 0.86 | 1 |

*95% CI: 95% confidence interval*

**Supplementary Table 3. Intraclass correlation coefficient (ICC), standard error of measurement (SEM) and Minimal Detectable Change (MDC) at the 90% level (MDC_90_). Values are shown separately for men and women.**

| **Test** | **Cohort** | **Valid tests** | **ICC** | **ICC 95% CI** | **SEM** | **MDC_90_** | **Rounded MDC_90_** |
| --- | --- | --- | --- | --- | --- | --- | --- |
| Immediate Memory | Entire Cohort | 16,612 | 0.743 | [0.734, 0.753] | 1.668 | 3.881 | 4 |
|  | Men | 12,712 | 0.738 | [0.727, 0.749] | 1.675 | 3.897 | 4 |
|  | Women | 3,900 | 0.758 | [0.739, 0.777] | 1.646 | 3.829 | 4 |
| Delayed Recall | Entire Cohort | 16,612 | 0.766 | [0.757, 0.775] | 0.813 | 1.891 | 2 |
|  | Men | 12,712 | 0.754 | [0.743, 0.765] | 0.831 | 1.934 | 2 |
|  | Women | 3,900 | 0.801 | [0.785, 0.817] | 0.757 | 1.761 | 2 |
| Digits Backwards | Entire Cohort | 21,090 | 0.694 | [0.684, 0.704] | 0.446 | 1.037 | 1 |
|  | Men | 16,710 | 0.681 | [0.669, 0.692] | 0.447 | 1.04 | 1 |
|  | Women | 4,380 | 0.733 | [0.714, 0.753] | 0.442 | 1.029 | 1 |
| Concentration | Entire Cohort | 21,087 | 0.688 | [0.678, 0.698] | 0.465 | 1.082 | 1 |
|  | Men | 16,707 | 0.677 | [0.665, 0.688] | 0.465 | 1.083 | 1 |
|  | Women | 4,380 | 0.722 | [0.702, 0.742] | 0.463 | 1.076 | 1 |
| Single-leg stance | Entire Cohort | 21,091 | 0.652 | [0.641, 0.663] | 0.935 | 2.174 | 2 |
|  | Men | 16,710 | 0.651 | [0.638, 0.663] | 0.929 | 2.161 | 2 |
|  | Women | 4,381 | 0.654 | [0.63, 0.679] | 0.957 | 2.225 | 2 |
| Tandem stance | Entire Cohort | 21,091 | 0.806 | [0.800, 0.812] | 0.461 | 1.073 | 1 |
|  | Men | 16,710 | 0.802 | [0.795, 0.809] | 0.467 | 1.088 | 1 |
|  | Women | 4,381 | 0.823 | [0.810, 0.835] | 0.435 | 1.013 | 1 |

*95% CI: 95% confidence interval*
